# CHANGER: A novel genetic reference from 902 unrelated Chilean exomes to enhance South American medical genetics and research

**DOI:** 10.64898/2026.01.27.26344982

**Authors:** Evelin González, Camilo Villamán, Boris Rebolledo-Jaramillo, Carlos F. Hernandez, Bernabé I. Bustos, Dominga Berrios, Gabriela Moreno, Jennifer E Posey, James R. Lupski, Cecilia Poli, Juan Francisco Calderón, Paula Muñoz-Venturelli, Mario I. Fernández, Juan Alberto Lecaros, Ricardo Armisén, Gabriela M. Repetto, Eduardo Pérez-Palma

## Abstract

**Background:** Genetic studies have disproportionately focused on populations of European ancestry, limiting the generalizability of allele-frequency references and genetic associations to underrepresented groups, including South American populations. This gap is particularly relevant for rare diseases and cancer, where accurate variant interpretation depends in part on appropriate population context. In addition, population-specific haplotype structure influence genome-wide association analyses and the portability of polygenic scores across ancestries. By focusing on the Chilean population, our work aims to bridge these gaps, providing more accurate reference data for genetic research and enhancing diagnostic and therapeutic strategies for underrepresented communities.

**Methods:** We aggregated exomes from seven Chilean cohorts and processed samples using a standardized best practices workflow, including population-scale quality control and relatedness filtering. Aggregated variants were annotated with VEP (including LOFTEE, REVEL, AlphaMissense, EVE, and CADD) and clinically classified using InterVar. Population structure was assessed with PCA/admixture. We compared overlap and allele frequencies against external references, performed gene-level variant burden analysis, and constructed phased haplotypes for ancestry association using linear regression with Bonferroni correction. Finally, we developed a dashboard with R shiny framework to enable gene-wise exploration of annotated variants.

**Results:** We present CHANGER (Chilean Aggregated National Genomics Resource), a comprehensive aggregation of 902 unrelated Chilean exomes designed to create a detailed reference of genetic variation in Chile. By incorporating data from multiple cohorts, we identified 774,110 unique genetic variants, with an average of 42,601 aggregated variants per individual. We identified 132,363 novel variants, of which 31,470 were common within our cohort (frequency > 1 %). We provide variant annotation and direct comparisons with other publicly available general population references. Beyond variant discovery, CHANGER supports gene-level burden scans (61 Missense-Damaging depleted genes; 2 Loss-of-Function enriched genes) and a haplotype resource for ancestry association (51,171 haplotypes), enabling downstream interpretation, improved imputation, and more powerful genome-wide association analyses in Chileans.

**Conclusions:** CHANGER provides a valuable resource for genetic research and a reference for local variant interpretation. Importantly, CHANGER follows a high-quality methodological baseline and an open, FAIR-aligned infrastructure designed to grow, increasing its value for discovery, imputation, and equitable clinical interpretation over time.

## Background

The widespread adoption of next-generation sequencing has reshaped human genetics by enabling systematic catalogues of both normal and pathogenic variation. Reference resources such as the 1000 Genomes Project(1), the Human Genome Diversity Project (HGDP)(2), and gnomAD(3) have been foundational in establishing allele-frequency priors, constraint metrics, and variant interpretation frameworks.(2–4) Yet these resources remain disproportionately represented by samples of European ancestry, reflecting long-standing imbalances in sampling(5,6). The result is an incomplete representation of global diversity, with immediate consequences for both research and clinical practice in underrepresented populations.(7)

Variant interpretation is directly affected: alleles that are common or benign in an underrepresented ancestry may appear “rare” or “pathogenic” in Eurocentric databases and thus be misclassified elsewhere.(7,8) The clinical implications are well illustrated by examples such as cardiomyopathy genetics, where false “pathogenic” assignments were disproportionately returned to African American patients; reanalysis with ancestry-appropriate controls corrected many of these errors(9). Rare disease studies in underrepresented populations are similarly affected. For example, in a UK large scale study, probands with African ancestry were less likely to receive a diagnosis(10). Moreover, hidden bottlenecks remain unexplored: founder mutations in understudied populations, such as the *COL27A1* variant causing Steel syndrome in Puerto Ricans, highlight how pathogenic alleles can persist undetected for decades when local references are missing(11). Similarly, genome-wide association studies in Chileans have revealed risk *loci*, such as *TRAF3* for gallstone disease, that were absent or only weakly detected in Europeans, highlighting the discovery potential of ancestry-specific signals(12). Complex trait genetics is equally constrained. Polygenic risk scores (PRS) derived from European discovery cohorts show markedly reduced predictive power in other populations, with prediction accuracy falling by up to 4.5-fold in Africans and ∼1.6-fold in Latinos(5). Simulations confirm that the correlation between true and inferred genetic risk decays with increasing genetic divergence from the discovery population, even when causal variants are shared(6). Without systematic inclusion of diverse cohorts, PRS and similar tools risk exacerbating existing health inequities.(5,7) Somatic oncology is also impacted, as germline background frequency priors are essential to filter tumor–normal comparisons, and Eurocentric references reduce the specificity of variant calls in admixed or non-European patients.(8)

Among global datasets, gnomAD v4.1 is the most comprehensive reference for general population variation, aggregating more than 800,000 individuals (exomes and genomes combined) across multiple ancestries.(3,13) It provides allele-frequency estimates, mutational constraint metrics, and joint counts that inform both research and clinical workflows. Clinically, gnomAD frequencies can be integrated into the American College of Medical Genetics and Genomics (ACMG) guidelines for variant interpretation (e.g. BA1, BS1 and PM2 thresholds)(4), while constraint informs candidate gene interpretation. However, despite its scale, ancestry representation remains uneven: European (EUR) samples dominate, while Admixed Americans (AMR), particularly South American populations remain underrepresented.(6) Although the most recent version gnomAD (v4.1) increased by 1.7x the amount of Admixed Ameridian individuals (AMR), this is still an insufficient improvement, especially when considering that 77% of the data in gnomAD belongs to EUR population, and only 3.72% belongs to AMR.(13)

Admixed South Americans populations are shaped by diverse indigenous groups, European colonization, and forced African migration(14). This yields haplotype mosaics and allele-frequency distributions that diverge from European populations. Local-ancestry inference in admixed cohorts has shown that Amerindian tracts are under-supported by reference haplotypes, leading to lower imputation accuracy and reduced coverage at clinically relevant loci(6,15). This in turn limits the power in detecting ancestry specific signals in genome-wide association studies (GWAS). Current data are scattered, often limited to specific cohorts, and not optimized for clinical interpretation. Without ancestry-matched controls or general population references, laboratories risk overcalling variants as causal or undercalling those that are truly pathogenic, particularly in *de novo*-enriched disorders such as neurodevelopmental syndromes or other rare diseases. The result is an elevated burden of variants of uncertain significance (VUS) and prolonged diagnostic odysseys in underrepresented groups.(7,14)

The inequities in representation are recognized within the region. Several national initiatives aim to build resources tailored to local diversity. The Mexican Biobank of 6,057 individuals across all 32 states(16) and genomic data from ∼140,000 adults in Mexico City(17) lead these efforts in the region. Argentina’s PoblAr biobank is designed to capture the breadth of the country’s genetic variation^18^. Brazil’s BIPMed and ABraOM (Brazilian genomic variants) initiatives promote genomic data sharing and implementation in public health(18,19) and have evolved into a country-level GenomasBrazil project, funded by the National Health System (SUS)(20). Colombia has piloted whole-genome sequencing for rare-disease diagnostics(21) and The Consortium for Genomic Diversity, Ancestry, and Health in Colombia (CÓDIGO)(22). Chile has characterized a small number of native Mapuche Huilliche individuals(23) and launched ChileGenómico, focusing on characterizing national diversity rather than focusing on human genetics(24).

Chile exemplifies these challenges. Its demographic history reveals sex-biased admixture (European male / Native American female), a north–south gradient of African ancestry, and substantial regional heterogeneity in Indigenous contributions, particularly Mapuche ancestry in the south(23,25). Central Chile, home to the majority of the population, assimilated the largest wave of European immigration, producing distinct geographic gradients in allele frequencies(25). These features mean that classical pan-Latin American references such as AMR, Native American (NAT) or gnomAD’s Latino subset are inadequate proxies for Chile(23).

There is a need for broader representation of population and genomic data. Here we present CHANGER (Chilean Aggregated National Genomics Resource), a comprehensive aggregation of 902 unrelated Chilean exomes designed to fill this gap.

## Methods

### Contributing projects

Data from seven different cohorts were aggregated for this initiative (Table 1). Briefly, the projects are as follows: 22q aimed at identifying genetic modifiers of the presence of congenital cardiac anomalies in individuals with 22q11.2 microdeletion(26). AORTIC-Chile samples were obtained from the “Registry of Chilean Patients with Connective Tissue Disorders” including patients with clinical diagnosis and compatible clinical manifestations.(27) Oncoprecisión Chile performed whole exome sequencing in tumors from individuals diagnosed with urothelial bladder cancer from arsenic-exposed and non-exposed regions in Chile (Antofagasta and Santiago, respectively). DECIPHERD performed trio exome sequencing for patients with undiagnosed rare diseases, presenting with multiple congenital anomalies, intellectual disability and/or immune disorders; for this aggregation, unaffected parents were also included(28).

**Table 1.** CHANGER contributing datasets.

| Project ID | Phenotype in Study | Method | Affected Individuals | Unaffected individuals | Total | Bioethical approval ID |
| --- | --- | --- | --- | --- | --- | --- |
| 22q | 22q11.2 microdeletion syndrome | Exome | 55 | 0 | 55 | CEC-UDD 2012-20 |
| AORTIC CHILE | Marfan Syndrome and connective tissue disorders | Exome | 13 | 20 | 33 | CEC-UDD 2018-61 |
| Oncoprecision Chile | Non-familial Bladder Cancer | Exome | 35 | 28 | 63 | CEC-UDD 2022-09 |
| DECIPHERD | Undiagnosed rare diseases | Exome | 8 | 172 | 180 | CEC-UDD 2018-045 |
| FONEPI | Epilepsy (GGEs, DEEs, other) | BGE | 37 | 430 | 467 | CEC-UDD 2022-21 |
| PIGIT | Inborn Errors of Immunity | Exome | 10 | 70 | 80 | CMG/GREGoR H-29697 (BCM-IRB) |
| Rett | Rett and Rett-like syndromes | Exome | 0 | 24 | 24 | CEC-UDD 2014-113 |
| **Total** |  |  |  |  | **902** |  |

*GGE: Genetic Generalized Epilepsies; DEE: Developmental and Epileptic Encephalopathies; BGE: Blended Genome Exome. CEC-UDD: Comité-Ético-Científico (Ethical and Scientific Committee) Universidad del Desarrollo.*

FONEPI cohort, an ongoing nationwide effort, aims to identify how common and rare genetic variants shape disease risk and presentation. It includes genetic generalized epilepsy cases, general population controls and trios with developmental epileptic encephalopathies. For the latter, only unaffected parents were included. The Program of Immunogenetics and Translational Immunology (PIGIT) performed mostly trio exome sequencing of patients with suspected inborn errors of immunity. Rett is a genomic analysis project for patients with a suspected clinical diagnosis of Rett syndrome.(29)

### Ethical approval

The CHANGER aggregation project was reviewed and approved by the Institutional Review Board (IRB) of the Facultad de Medicina, Clínica Alemana–Universidad del Desarrollo *(Comité-Ético-Científico*, CEC-UDD). The same IRB had previously approved each dataset contributing to the aggregation. All participants provided written informed consent, including permission to use their DNA in genetic studies and to share aggregated genomic data.

### Variant calling workflow

In total, 1,106 exome samples were compiled from available datasets. All samples were processed using the multi-step GATK Best Practices pipeline for germline variant calling, following the tutorial by Sealock *et al*(30), using the publicly available references from the Broad Institute. Briefly, the analysis relied on Picard v2.26.10, GATK v4.6.1.0(31), and Samtools v1.11, with default parameters unless otherwise specified. The raw data consisted of three formats: FASTQ (n=510), BAM (n=174), and CRAM (n=422). FASTQ and BAM files were converted to unmapped BAMs (uBAMs) so that all samples could be processed identically, while CRAM files entered the pipeline directly. All reads were aligned to the hg38 reference genome with BWA-MEM v0.7.15. Initial read-level QC metrics were collected with Picard CollectMultipleMetrics, including base composition by cycle, insert size distribution, mean quality by cycle, and quality score distribution. We carried out sorting, recalibration and validation with Picard SortSam, GATK BaseRecalibrator and Picard ValidateSamFile, ensuring file integrity before downstream analysis(31). Per-sample variant calls were generated with GATK HaplotypeCaller. Resulting gVCFs were reblocked with GATK ReblockGVCF.

### Sample aggregation

Multi-sample VCFs were then aggregated and processed with Hail v0.2, applying filters described by Sealock *et al.*(30). These included thresholds for read depth (DP=10), genotype quality (GQ=20), allelic balance (AI=0.2), and call rate (CR=0.95), balancing sensitivity (capturing true rare variants) with specificity (excluding artifacts) across hundreds of exomes. Cryptic relatedness was evaluated with the Hail implementation for the kinship coefficient(32). When two samples showed a kinship >0.2 (consistent with second-degree relatives), the individual with fewer called variants was excluded. This ensures that the final dataset consists of unrelated individuals.

### Population structure analysis

To characterize ancestry, we performed principal component analysis (PCA) and admixture analysis. PCA provides a global view of genetic structure by summarizing genome-wide variation into axes of differentiation, while admixture analysis estimates the proportions of ancestry components per individual. We constructed a comprehensive reference panel by combining all individuals from the 1000 Genomes Project (1KGP) and the Human Genome Diversity Project (HGDP)(2) with the Chilean Native (HUI) samples previously characterized by Vidal et al. (2019)(23). PCA was performed following the methodological approach described by Leal et al. 2025(33), using HUI, AMR, AFR and EUR as reference populations. Global ancestry proportions were estimated using a supervised framework implemented in ADMIXTURE(33,34). We used a subset of non-admixed samples described by Shriner *et al*.(35) as reference individuals. Ancestry proportions were inferred under two models: one including three continental populations (AMR, AFR, EUR, and another including four populations (HUI, AMR, AFR and EUR).

### Variant annotation

Variants were annotated with Variant Effect Predictor (VEP), using databases and predicting algorithms including gnomAD v4.1.1(13), EVE(36), AlphaMissense(37), REVEL(38), LOFTEE(3) and CADD(39). Variant-level annotations (REVEL, CADD_PHRED, AlphaMissense am_class, EVE EVE_CLASS, LOFTEE LoF) were parsed from MANE-select per-gene transcript. A variant was flagged as “Damaging” if any criterion was met: REVEL ≥ 0.75; CADD_PHRED ≥ 20; AlphaMissense (am_class) = pathogenic or likely_pathogenic; EVE EVE_CLASS = pathogenic or likely pathogenic; or LOFTEE LoF = “HC” (high-confidence LoF). For clinical interpretation, we applied InterVar(40), which semi-automates ACMG/AMP guidelines to classify variants as pathogenic, likely pathogenic, uncertain, likely benign, or benign.

### Gene-wise variant burden Analysis

We tested damaging-missense (Miss-D) to synonymous (SYN) and loss-of-function(LoF) to synonymous per gene rates in CHANGER (CH) against gnomAD. To obtain gnomAD Miss-D and SYN counts, we implemented the aforementioned CHANGER VEP annotation pipeline and damaging criteria on available VCFs in gnomAD webpage (https://gnomad.broadinstitute.org/data)(13). For LoF variant analysis we used gnomAD LoF and SYN counts (lof.obs, syn.obs) from constraint metrics, also available in the gnomAD database. We applied a two-sided Fisher’s exact test. We called significant outliers by a 10x rule: a gene was considered variant-enriched if Benjamini-Hochberg (BH) FDR≤0.05 & log₂OR>log₂10 or variant-depleted if BH FDR≤0.05 & log₂OR<−log₂10. For both analyzes, we enforced a stability filter requiring genes to have CHANGER SYN≥10 and gnomAD SYN≥100. To avoid noise we excluded genes associated with exome sequencing false-positive signals reported in Fajardo K, *et al*(41).

### Haplotype-based analysis

LD-based haplotype blocks were called on autosomes and chrX using common SNPs (MAF ≥1%) with the ‘blocks’ function in PLINK v1.9 (default parameters)(42). Within each block, phased genotypes were obtained from chromosome-wise phasing of the Chilean exomes VCFs with SHAPEIT5 v1.0(43), using genetic maps in the human reference genome version GRCh38 and processing chromosome X in two parts: (i) a non-pseudoautosomal (non-PAR) run restricted to XX samples, and (ii) a PAR-specific run including all XX and XY samples together. Using custom Python scripts, we combined the PLINK block definitions with the SHAPEIT5-phased BCFs (binary VCFs) to identify allele order of haplotypes, encode the SNPs alleles within the blocks (0/1), set the allele sequence, and compute per-sample haplotype dosages (0/1/2 copies). These dosage matrices were then used merged with the most significant individual-level ancestry proportions (EUR, AMR, HUI) along with covariates (study cohort for all chromosomes, plus sex for chrX), and for each haplotype block we fit a linear regression: ancestry_trait ∼ haplotype_dosage + sex + study_cohort in R (glm function), applying a minimal allele count (MAC) threshold of ≥20 on autosomes and a MAC ≥11 for chrX non-PAR in chromosome XX samples.

Association results were shown as exome-wide Manhattan and QQ plots, using Bonferroni correction for multiple testing (p = 9.77e-07). Haplotype blocks were annotated to overlapping genes using gene definitions available in GENCODE v.48(44). Finally, we visualized the strongest ancestry-associated haplotype blocks as a heatmap of a combination of the -log10 p values and beta coefficients (-log10P x sign(Beta)) across EUR, AMR and HUI ancestry components.

### CHANGER dashboard

We developed a dashboard web application using the R Shiny framework with a server-side reactive pipeline, as previously reported(45,46). Variant annotations (e.g., VEP fields, LOFTEE, REVEL, CADD, AlphaMissense, EVE, gnomAD joint metrics) and MANE-select transcript metadata (strand, coding exons, CDS length, block sizes) are integrated, and kept unmodified from source(47). For deployment efficiency, per-gene tables with main counts and annotations were consolidated into a single SQLite database using DBI/RSQLite with an index on gene symbols. Flags are computed as: Damaging as previously defined, LoF=LOFTEE High Confidence; Common if AC≥9; CHANGER_AF as AC/AN. A strand-aware parser maps HGVSc cDNA positions to transcript exonic coordinates in a lollipop format with LoF, Missense and Synonymous variants denoted by color. For simplicity we mapped intronic, upstream, and downstream variants between exons (or before and after when corresponds) in a predefined standard space within the transcript plot (10% length in total). We included external variant links to gnomAD if the variant is previously reported. All visualizations use ggplot2 library. Deployment to shinyapps.io was done via rsconnect library. Code for dashboard is available at: https://github.com/edoper/changer.

## Results

### Sample Aggregation

We aggregated a total of 1,106 exome samples from seven cohorts, of which 902 unrelated individuals passed quality control and relatedness filtering. Each contributing cohort’s main features and number of participants in CHANGER are shown in Table 1. For 727 aggregated individuals (80.6%), both municipality and region of origin were available, covering 114 of Chile’s 346 municipalities and all 16 regions (Figure 1A). Regional distribution of participants was correlated with regional population density (Supplementary Figure 1, Pearson r = 0.97, p = 4.1×10^-10^)(48). After standardized pre-processing, alignment, and base recalibration, the final callset comprised 38,426,324 alleles aggregated into 774,110 unique high-quality variants (Supplementary Figure 3). Across individuals, the mean variant count was 42,601 per exome, consistent with expectations for aggregation of high-coverage human exomes(49). Across the 774,110 variants in the dataset, 391,891 were singletons, carried by only one individual. In contrast, 153,461 variants (19.8%) were common, present in at least 1% of the cohort (≥9 carriers). The site frequency spectrum showed an excess of rare variants, with 50.6% of all variants present at <0.1% frequency, mirroring patterns reported in large-scale exome projects (Figure 1B)(3,49).

**Figure 1.**
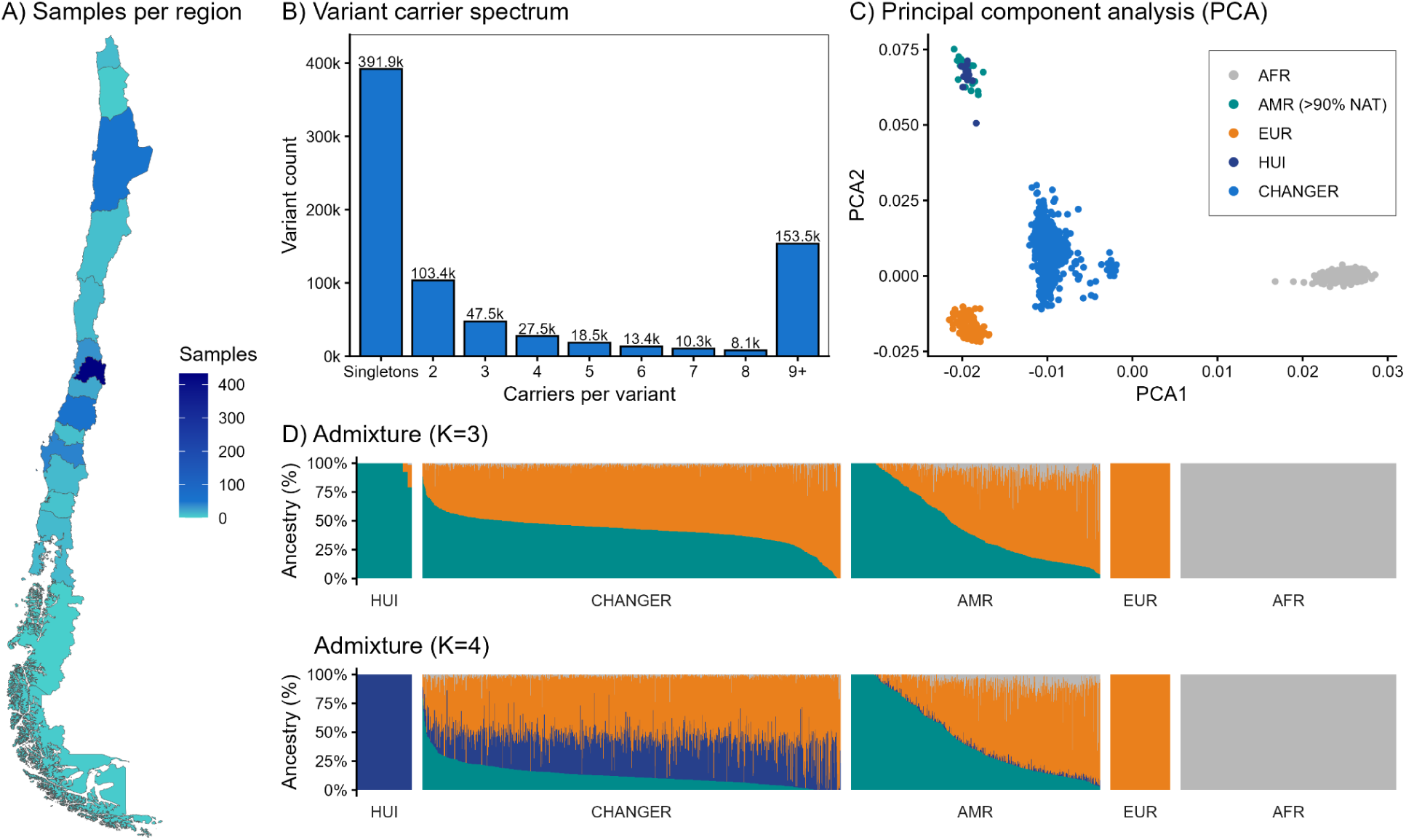
CHANGER aggregated individuals, Allele frequency spectrum and population structure. A) Choropleth map of Chile showing CHANGER individuals per region (continental Chile). We extracted municipality and regional Polygons from the Chilemapas package v3 (https://pacha.dev/chilemapas/. B) Variant carrier spectrum with variants binned by number of carrier individuals; bar tops display total counts and the y-axis is shown in thousands. C) Principal component analysis of CHANGER (n=902) overlaid on non-admixed reference panels: AFR in gray (n = 471), EUR in orange (n = 136), HUI in dark blue (n = 12) and AMR in dark green (n = 23). These 23 AMR individuals correspond to a subset with >90% NAT ancestry. D) Top panel: Admixture proportions at K = 3 across groups (HUI = 12, CHANGER = 902, AMR = 538, EUR = 136 and AFR = 471, individuals ordered within facets. Bottom panel: Admixture proportions at K=4 across the same groups. Coloring of ancestry proportions remains as in C), that is, gray, orange, dark blue and dark green for AFR, EUR, HUI and AMR (NAT) ancestries. Although the ADMIXTURE model was trained using the non-admixed AMR subset (n = 23), the full AMR population (n = 538) is displayed to illustrate the distribution pattern relative to CHANGER.

### Allele frequency spectrum and population structure

Principal component analysis (PCA) positions the CHANGER cohort along the EUR–NAT axis with minor contribution of the African (AFR) cluster (PC1 = 56.14%, PC2 = 16.50% of variance), with Huilliche (HUI, n = 12) and admixed Ameridian individuals (AMR >90%NAT, n = 23) populating a NAT cluster (Fig. 1C). CHANGER PC1 vs PC2 distribution is consistent with prior demographic studies of Chile(23,25) and with recent South American assessments using similar reference panels(33). Admixture at K=3 resolves continental components in CHANGER (n = 902): 55.96 ± 12.50% EUR, 41.82 ± 12.68% NAT, and 2.23 ± 2.78% AFR. At K = 4, the NAT ancestry partitions into a HUI-NAT component and a broader AMR-NAT component; CHANGER averages 31.16 ± 12.26% HUI-NAT, 12.86 ± 10.09% AMR-NAT, 54.16 ± 12.83% EUR, and 1.82 ± 2.81% AFR, totaling 44.02 ± 13.14% NAT (HUI + AMR). In contrast, the AMR reference panel (n = 538; individuals largely from the US, Central America, Colombia, and Peru) exhibits HUI-NAT 1.76 ± 2.15%, with NAT ancestry dominated by the AMR-NAT component (44.74 ± 32.11%; European 46.49 ± 28.13%, African 7.01 ± 8.38%). Interestingly, modeling the HUI-NAT component at K = 4 increases estimated NAT-like ancestry in CHANGER by ∼2.20 percentage points relative to K = 3. A Wilcoxon rank-sum test comparing K = 4 HUI-NAT ancestry between CHANGER and AMR is highly significant (U = 473,430; p = 1.65×10⁻²⁰¹), indicating that Native-American ancestry is specifically enriched for a Huilliche-like component that remains marginal in broader American references (Figure 1D).

### Comparative analysis with global references

The intersection analysis between the CHANGER dataset and major global reference resources (Figure 2A) revealed that, among the 774,110 variants identified in CHANGER, 639,490 (82.6%) were also present in gnomAD(13), 352,503 (45.5%) in ABraOM(19), and 341,196 (44.1%) in HGDP/1000G(2). The number of variants shared across all three population datasets reached 288,831 (37.3%), underscoring a substantial overlap with worldwide genomic resources. In contrast, 132,363 variants (17.1%) were unique to CHANGER, not represented in any of the three external datasets. Within this CHANGER-specific fraction, 31,470 variants (4.1% of all CHANGER variants; 23,7% of all unique CHANGER variants) exhibited a minor allele frequency ≥ 1% within the Chilean cohort yet were absent from gnomAD, HGDP/1000G, and ABraOM. These represent variants that are common within the local population but unrecorded globally, highlighting the importance of incorporating population-specific data to capture allelic diversity that remains unrepresented in international repositories. The total variant counts of the comparison datasets spanned from 183.7 million in gnomAD, 79.9 million in HGDP/1000G, and 75.9 million in ABraOM (Figure 2A). While CHANGER contained fewer variants overall due to its exome-based design and more limited sample size, it contributed to a distinct subset of frequent regional alleles that expand current catalogs of human genomic variation. Using the full intersected variant set, we compared variant-level AF values against gnomAD v4.1 (n = 807,162), gnomAD-AMR subset (n = 30,019), and the Brazilian AbraOM (n = 1,171) individuals. Overall, allele frequencies in CHANGER showed strong linear correlations with all three datasets, reflecting the shared continental ancestry structure. However, a subset of variants showed significant frequency discrepancies. After Bonferroni correction, 3,576 variants in gnomAD, 3,123 in gnomAD-AMR subset, and 2,595 in AbraOM were identified as significant outliers (standardized residuals > 3). These variants represent alleles whose frequencies in Chile differ significantly from global and regional references, highlighting population-specific frequency shifts within South America. Across all individuals, the mean number of novel variants was 706.6 (95% CI 694.2–718.9). When comparing individuals at the tails of NAT ancestry (K3), the number of novel variants in individuals at the top 20% of NAT ancestry (n= 705.3, 95% CI 683.3–727.4) was significantly higher than the novel variants observed in individuals at the bottom 20% (n= 690.8, 95% CI 673.1–708.5, Wilcoxon rank-sum test p = 0.025).

**Figure 2.**
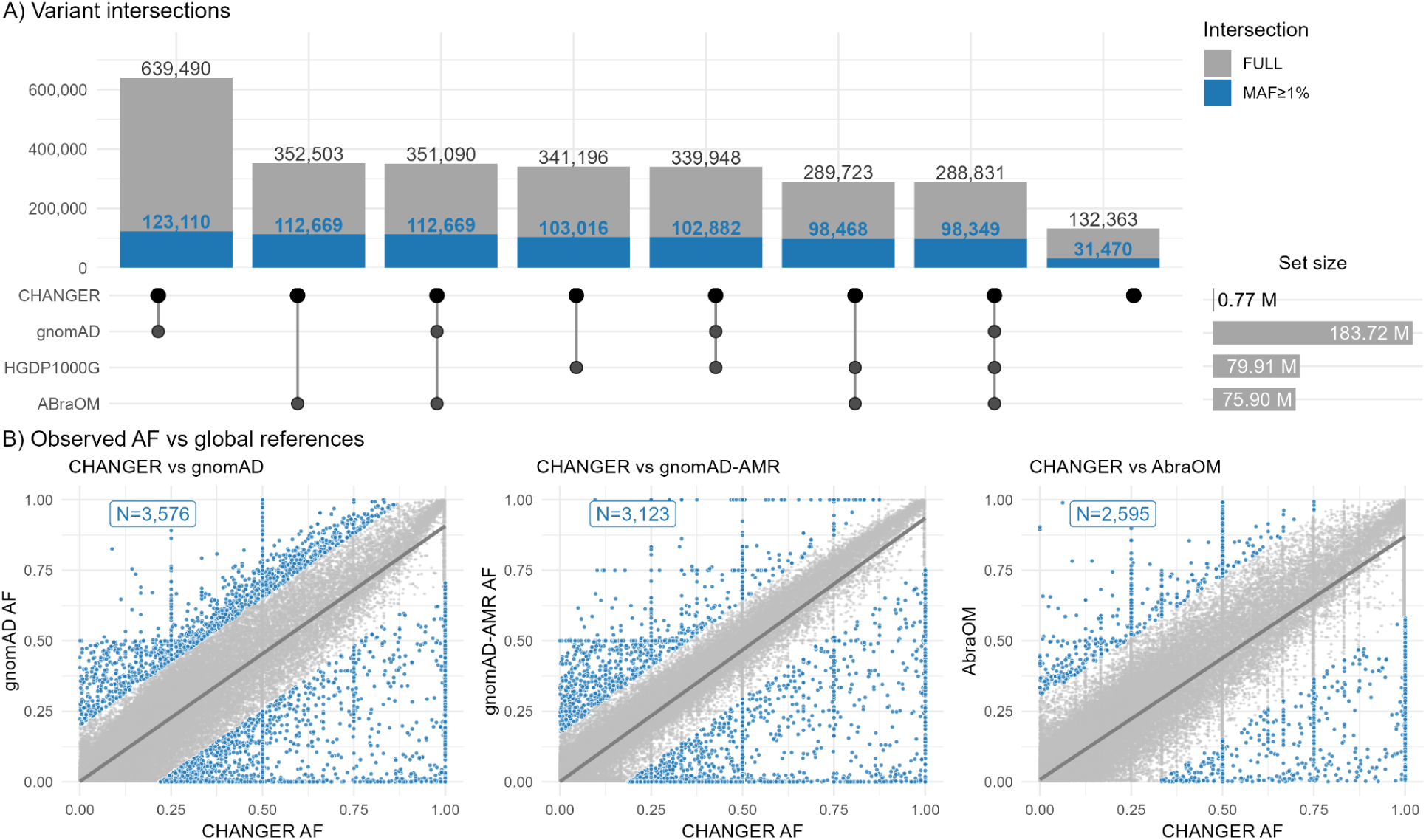
Changer variant intersections across genomic datasets. A) Intersection plot showing the number of genetic variants shared between the CHANGER dataset and major population resources (GnomAD, HGDP/1000G, and ABraOM). Bars represent the total count of variants shared between (grey) and major population resources. We highlight CHANGER common variants (MAF ≥1%) in blue across each intersection. The lower panel depicts the corresponding set combinations included in each intersection. Total number of variants per intersected dataset (Set size), expressed in millions is shown in horizontal bars. B) Correlation between CHANGER observed allele frequencies (AF) and reported AF on gnomAD, gnomAD-AMR subset and AbraOM on corresponding intersected variants. CHANGER variants with significantly different AF are shown in blue (>3SD, Bonferroni adjusted p-value < 0.05) with the corresponding total number of significant variants shown in blue box (upper left corner).

### Pathogenicity analysis

A total of 1,073 variants were annotated as pathogenic or likely pathogenic according to InterVar. Among these, we highlight two variants with high allele counts in CHANGER but very low or absent frequencies in global references: one in the *PDPR* gene (NM_001322117.1:c.T182A p.L61H), with AC = 0 in gnomAD, AC = 49 (1 case and 48 controls) in CHANGER, and one in the *CACNA1C* gene (NM_001129837.1:c.G6013T p.E2005X), with AC = 1 in gnomAD, AC = 13 (all controls) in CHANGER. The *PDPR* gene (OMIM* 617835) has not been associated with a disease according to OMIM, although it has been proposed as a candidate for neurodevelopmental disorders(50) and developmental delay with brain anomalies(51), whereas *CACNA1C* (OMIM *114205) has been associated with dominant forms of Brugada Syndrome 3, Long QT syndrome 8, Neurodevelopmental disorder with hypotonia, language delay, and skeletal defects with or without seizures, and Timothy syndrome.

### Gene-wise variant burden analysis

We searched for genes that are depleted or enriched for variation in CHANGER relative to gnomAD. We interrogated two classes of variants: missense-damaging (Miss-D) and high-confidence loss-of-function (LoF) variants. Using per-gene variant class-to-synonymous rates for normalization and a BH FDR < 0.05 multiple testing correction, a gene was only considered significant if in addition it was 10 times depleted or enriched in comparison to gnomAD variant counts (See Methods). We observed a set of 61 genes with significant depletions of missense-damaging variants (Figure 3A, Supplementary Table 1); among these we highlight the ten most depleted genes (*TBC1D15, WNT7B, ARHGAP35, SLC43A2, FBXW9, RB1CC1, ANKRD17, GET4, MAEA and STT3A)*. We observed no enrichments for missense-damaging variants in CHANGER relative to gnomAD. In contrast, for LoF high confidence variants, applying the same framework yielded two genes with significant 10-fold enrichments (Figure 3B, *PRR36*, OR = 16.18 and *SON*, OR = 51.54). No significant LoF depletions were observed. Together, these analyses indicate that, even at this sample size and under conservative calling and effect-size thresholds, strong outliers (depleted and enriched) can be identified in CHANGER.

**Figure 3.**
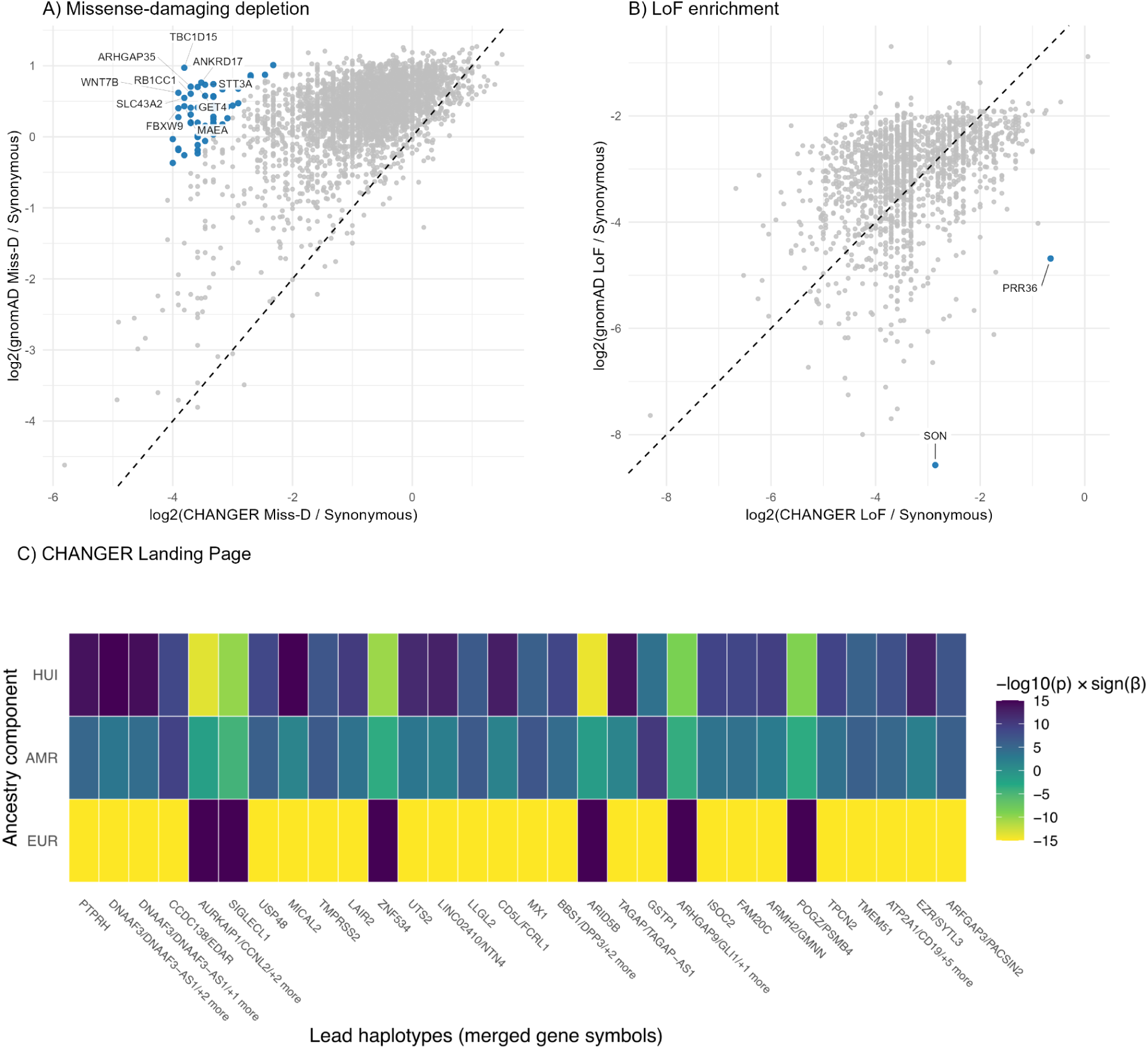
Variant burden and haplotype-based analysis. A) Gene-wise missense-damaging (Miss-D) burden analysis. Scatter of log₂ rates Miss-D/Synonymous in CHANGER (x-axis) versus gnomAD (y-axis) are shown for every gene included in the analysis (n=3,497). Significantly depleted genes with at least 10x effect (n=61) are shown in blue. Top 10 genes with the highest observed depletion are labeled. B) Gene-wise Loss-of-Function (LoF) burden analysis. Scatter of log₂ rates LoF/Synonymous in CHANGER (x-axis) versus gnomAD (y-axis) are shown for every gene included in the analysis (n=1,627). Significantly enriched genes with at least 10x effect (n=2) are labeled and shown in blue. For A and B, dashed diagonal is y=x. C). Top 30 significant haplotypes associated with ancestry components. Each column represents a lead haplotype mapped to one or more genes, and each row shows one ancestry component (EUR, AMR, HUI). Colors indicate the direction and strength of association (–log₁₀(p) × sign(β); scale –15 to 15), with dark purple indicating higher haplotype dosage in individuals with higher ancestry values and yellow indicating the opposite. Haplotypes are sorted from higher to lower significance in a left to right manner.

### Haplotype-based analysis

We used LD-based blocks and SHAPEIT5 phasing to construct an exome-wide panel of 51,171 distinct haplotypes for 902 Chilean exomes (Supplementary Table 2). After Bonferroni correction at α = 0.05 (p < 9.77×10⁻⁷), we identified through linear regression 2,448 haplotypes significantly associated with EUR global ancestry proportions (Supplementary Figure 4A), 119 with AMR (Supplementary Figure 4B), and 731 with HUI (Supplementary Figure 4C). QQ plots revealed marked deviations from the null with genomic inflation factors ∼5.95 (EUR), ∼2.27 (AMR), and ∼3.92 (HUI) (Supplementary Figure 4 B, D, F), consistent with the fact that ancestry itself is a highly polygenic trait rather than a technical artefact. We next annotated each haplotype block to genes, resulting in 21,502 block–gene pairs and 15,816 genes with at least one overlapping haplotype (Supplementary Table 2). For each trait, we selected the most significantly associated haplotype per block and visualized the top 30 overlapping *loci* (Figure 3C). These top haplotypes show strong ancestry-specific effects, with some displaying associations to high levels of HUI/AMR ancestry (beta > 0) and low EUR ancestry (beta < 0; *PTPRH*, *DNAAF3/DNAAF3-AS1*, and *CCDC138/EDAR*) and others with the opposite effect (*AURKAIP1/CCNL2/+2*, and *SIGLECL1*). Together, these results show that the Chilean exome reference captures significant, ancestry-specific haplotypes.

### CHANGER Dashboard

To enhance the use of CHANGER in South American medical genetics research and variant interpretation, we developed a gene-centric (MANE-select transcripts) dashboard to explore variants aggregated (Supplementary Figure 5). The dashboard is available at: https://lngc.shinyapps.io/changer_v1/. For any queried gene, it presents summary metrics, an interactive lollipop map of variants across the transcript, and a scrollable variant table showing CHANGER allele frequency, “Damaging flag”, “Common (AF≥1%)”, and “Not in gnomAD” flags, with direct gnomAD links. Reactive filters (LoFTEE HC, Damaging, Not in gnomAD, Common) update visualizations and tables in real time. Here, gene-level aggregated variant tables are available for download.

## Discussion

Here, we present the very first exome-level variation database for an admixed Chilean population. CHANGER delivers variant annotation and direct comparisons with global references, offering ancestry-aware allele frequencies for both clinical interpretation and research design. CHANGER provides a population-anchored reference that improves rare-variant interpretation, strengthens local discovery, and contributes to a more equitable genomic landscape for South America.

A large number of reported variants were not present in global databases; thus, CHANGER contributes to improving knowledge about genomic variation. We identified 132,363 previously unreported variants. Of note, about 25% of unique CHANGER variants had allele frequencies over 1 % and thus are particularly common in the Chilean population.

From a population genetics perspective, CHANGER complements gnomAD and other international resources by anchoring analyses in a Chilean context using an established and comparative framework(30). Gene-level mutational constraint derived from gnomAD provides a global scaffold for variant interpretation^4^, but ancestry-specific frequencies are indispensable for interpreting local burden signals, enrichment analyses and discovery of population-private variants. As demonstrated in studies of admixed cohorts, local ancestry segments can carry unique alleles poorly represented in global datasets(15). CHANGER thus provides the necessary baseline for case–control designs, modifier analyses, and polygenic risk model calibration in Chileans, while also enhancing the comparability of regional studies within Latin America.

The creation of CHANGER directly addresses the diagnostic challenges faced by patients from underrepresented populations. In clinical genetics, the absence of ancestry-matched allele frequencies inflates the rate of variants of uncertain significance, delays diagnosis, and risks misclassification of benign or pathogenic variants(7,9). By providing a Chile-specific frequency baseline, CHANGER enables more accurate application of ACMG criteria, particularly for frequency-based evidence (e.g., BA1, BS1, PM2)(4,8). This not only reduces false-positive pathogenic assignments, as documented in cardiomyopathy studies with ancestry-imbalanced controls(52), but also improves confidence in variant classification and reclassification. Notably, *CACNA1C* results highlight the need for mechanism-aware interpretation: classic Timothy syndrome is associated with gain-of-function, whereas several other *CACNA1C* phenotypes (e.g., Brugada/short QT) are linked to loss-of-function. Given CACNA1C’s strong LoF constraint (gnomAD pLI≈1), the stop-gain p.E2005X observed is a useful discussion point, particularly in terms of whether it is predicted to undergo nonsense-mediated decay (vs escape NMD and produce a truncated protein)(53). In practice, the use of CHANGER should help shorten the diagnostic odyssey for families with rare disorders, a need already highlighted across Latin America(14).

Our database adheres to the FAIR principles (Findable, Accessible, Interoperable, Reusable), ensuring its long-term usability and integration into both clinical and research workflows(54). Public availability through our dashboard, clinicians are able to perform rapid allele lookups, while interoperability with standard formats and annotation frameworks facilitates incorporation into existing pipelines. This could enable CHANGER to participate in the future federated gnomAD initiative(55). Documentation of aggregation procedures and quality controls supports transparency and reuse(56). These features not only maximize the resource’s immediate impact but also position it for sustained contribution as sequencing technologies and analytic frameworks evolve.

To illustrate how CHANGER can be leveraged beyond allele-frequency lookup, we performed two complementary analyses: a gene-level depletion/enrichment scan against gnomAD and a haplotype-based association analysis with global ancestry components. Although the gene-level burden scan identified strong outliers—61 genes depleted for missense-damaging variation and two genes enriched for high-confidence LoF (*SON* and *PRR36*) relative to gnomAD (Figure 3A–B)—these findings should be interpreted cautiously given the current sample size (n=902). With this cohort size, extreme outliers can be sensitive to sampling variation, cohort composition, and residual annotation artefacts; replication in larger Chilean datasets and orthogonal biological/clinical validation will be important to confirm the most compelling signals. In contrast, the haplotype analysis leverages common-variant LD blocks and tests haplotypes under explicit MAC thresholds, yielding thousands of significant ancestry associations, and is therefore comparatively well powered at the current scale.

Complementing this, we inferred phased haplotypes and tested them for association with three EUR, AMR and HUI ancestry components. The resulting Manhattan and gene-level summaries show a highly polygenic architecture, with distinct haplotype effects on ancestry levels for the EUR component compared with the HUI and AMR components, and a clear differentiation between HUI and AMR, with haplotypes more specifically associated to indigenous ancestry of the Chilean population than general AMR ancestry. Similar population-specific reference panels, such as UK10K in the UK and the Haplotype Reference Consortium in Europeans, have shown that sequencing-based, population-matched haplotypes substantially improve imputation accuracy, particularly for low-frequency variants, and increase GWAS power(57,58), while multi-ancestry sequencing panels from TOPMed have likewise boosted imputation performance in African- and Hispanic-ancestry cohorts(59). Our phased haplotypes thus provide a valuable resource for building a Chilean-specific imputation panel, which may improve coverage of Chile-specific variants and increase power to discover novel signals in Chilean GWAS.

The multiple ethical and legal considerations for implementing responsible sharing of genomic data in CHANGER required reviewing and harmonizing with the ethical principles and guidelines of major international standards, and evaluating data access governance policy aligned with the new national legislation. This includes, in particular, the new personal data protection law (Law 21.719), which follows GDPR standards and comes into effect at the end of 2026, and the cybersecurity law in force since 2024 (Law 21.663)(60). Data were collected and stored in compliance with current national legislation on human research and personal data protection, through informed consent that authorizes sharing them aggregated and their future use with prior authorization from our IRB (CEC-UDD). We anticipate growth in this database and allowance for different access levels. For now, access is open to aggregated data, but later, once a full governance policy is implemented, controlled access through data access agreements for identified data could be feasible. We believe that standardizing data access processes overcomes one of the Latin America barriers to improving interoperability and fostering a data-sharing culture(61).

The limitations of this database include the fact that the data are derived from convenience samples rather than a random demographic design, and the relatively small sample size. Nevertheless, because the participating studies employed different ascertainment strategies, we demonstrated that the crowdsourcing strategy yielded adequate demographic representation across the country (Supplementary Figure 2). Strategies to expand the size and representation of this database will be crucial to enhancing its value and utility. Given that the samples originated from exome sequencing studies, non-coding and structural variation are evidently underrepresented, and closing these gaps will require further efforts in genome sequencing.

## Conclusions

Taken together, CHANGER demonstrates the value of national-level exome aggregation for underrepresented populations. By providing a clinically relevant, research-ready, and FAIR-aligned dataset, it enhances variant interpretation, empowers discovery, and reduces inequities in genomic medicine. More broadly, it illustrates a model for regional initiatives across Latin America, complementing global resources like gnomAD while ensuring that local demographic histories are adequately represented. As genomic medicine becomes increasingly central to healthcare, such efforts are essential to ensure that its benefits are distributed equitably.

## Ethics approval and consent to participate

This study was conducted in accordance with the Declaration of Helsinki and was approved by the relevant institutional ethics committees, including the corresponding Institutional Review Board of the Facultad de Medicina, Clínica Alemana–Universidad del Desarrollo (Comité Ético-Científico, CEC-UDD), Santiago, Chile. Written informed consent to participate was obtained from all participants (or their legal guardians with assent when applicable). Ethics approval IDs for the contributing cohorts are listed in Table 1 (including CEC-UDD approvals and BCM-IRB H-29697 for PIGIT/CMG-GREGoR).

## Consent for publication

Non applicable.

## Availability of data and materials

All data generated or analyzed during this study are included in this published article and its supplementary information files, including Figures 1–5 and Supplementary Table 1-2. Aggregated CHANGER data are publicly available through the CHANGER dashboard (https://lngc.shinyapps.io/changer_v1/), where gene-level aggregated variant tables can be downloaded. The dashboard source code is available at https://github.com/edoper/changer. Individual-level sequencing data are not publicly available due to informed-consent and privacy constraints.

## Author contributions

Conception and Design of the work: EPP, BRJ, RA, GMR,

Data acquisition EPP, BRJ, DB, GM, JP, JRL, CP, JFC, PMV, MIF, RA, GMR

Data analysis and interpretation. EPP, EG, CV, BRJ, CH, BIB, RA, GMR

Drafting and substantial revision of the manuscript EPP, BRJ, BIB, RA, GMR, JAL Approval of the submitted version; All Agreement to be both personally accountable for the author’s own contributions and to ensure that questions related to the accuracy or integrity of any part of the work, even ones in which the author was not personally involved, are appropriately investigated, resolved, and the resolution documented in the literature: All

## List of abbreviations

1KGP: 1000 Genomes Project
AC: Allele count
ACMG: American College of Medical Genetics and Genomics
AF: Allele frequency
AFR: African ancestry component
AMR: Admixed American ancestry component
AN: Allele number
BAM: Binary Alignment/Map (aligned reads file format)
BGE: Blended Genome–Exome sequencing
BH: Benjamini–Hochberg procedure (multiple-testing correction)
BWA-MEM: Burrows–Wheeler Aligner (MEM algorithm)
CADD: Combined Annotation Dependent Depletion (variant deleteriousness score)
CDS: Coding sequence
CEC-UDD: Comité Ético-Científico, Universidad del Desarrollo (ethics committee)
CHANGER: Chilean Aggregated National Genomics Resource
CI: Confidence interval
CR: Call rate (fraction of samples successfully genotyped at a site)
CRAM: CRAM (compressed alignment file format)
DEE: Developmental and epileptic encephalopathy(ies)
DP: Read depth (number of reads covering a genotype/site)
EUR: European ancestry component
EVE: Evolutionary model of Variant Effect (variant effect prediction score)
FAIR: Findable, Accessible, Interoperable, Reusable (data principles)
FASTQ: Sequencing read file format
FDR: False discovery rate
GATK: Genome Analysis Toolkit
GGE: Genetic generalized epilepsy/epilepsies
gnomAD: Genome Aggregation Database
gVCF: Genomic VCF (VCF recording per-site reference and variant blocks)
GRCh38: Genome Reference Consortium human build 38
GQ: Genotype quality
GWAS: Genome-wide association study
HGDP: Human Genome Diversity Project
HGVS: Human Genome Variation Society
HUI: Huilliche ancestry component (Mapuche subgroup)
INE: Instituto Nacional de Estadísticas (Chile)
IRB: Institutional Review Board
LD: Linkage disequilibrium
LoF: Loss-of-function
LOFTEE: Loss-Of-Function Transcript Effect Estimator (VEP plugin used by gnomAD)
MAF: Minor allele frequency
MANE: Matched Annotation from NCBI and EMBL-EBI
Miss-D: Missense-damaging
NAT: Native American ancestry component
NMD: Nonsense-mediated decay
OMIM: Online Mendelian Inheritance in Man
OR: Odds ratio
PCA: Principal component analysis
pLI: Probability of LoF intolerance
PRS: Polygenic risk score
QC: Quality control
QQ: Quantile–quantile (plot)
REVEL: Rare Exome Variant Ensemble Learner
SNV: Single nucleotide variant
SNP: Single nucleotide polymorphism
SQL: Structured Query Language
SYN: Synonymous (variant)
TOPMed: Trans-Omics for Precision Medicine
uBAM: Unmapped BAM
UTR: Untranslated region
VCF: Variant Call Format
VEP: Variant Effect Predictor
VUS: Variant of uncertain significance

## Data Availability

Aggregated CHANGER data are publicly available through the CHANGER dashboard (https://lngc.shinyapps.io/changer_v1/), where gene-level aggregated variant tables can be downloaded. The dashboard source code is available at https://github.com/edoper/changer. Individual-level sequencing data are not publicly available due to informed consent and privacy constraints. Additional summary data supporting the findings of this study are included in the published article and its supplementary information files.

## Acknowledgements

We thank each dataset participant who consented to participate in this research, and the clinicians who presented candidates to each included study.

## Supplementary Material

**Supplementary Figure 1.**
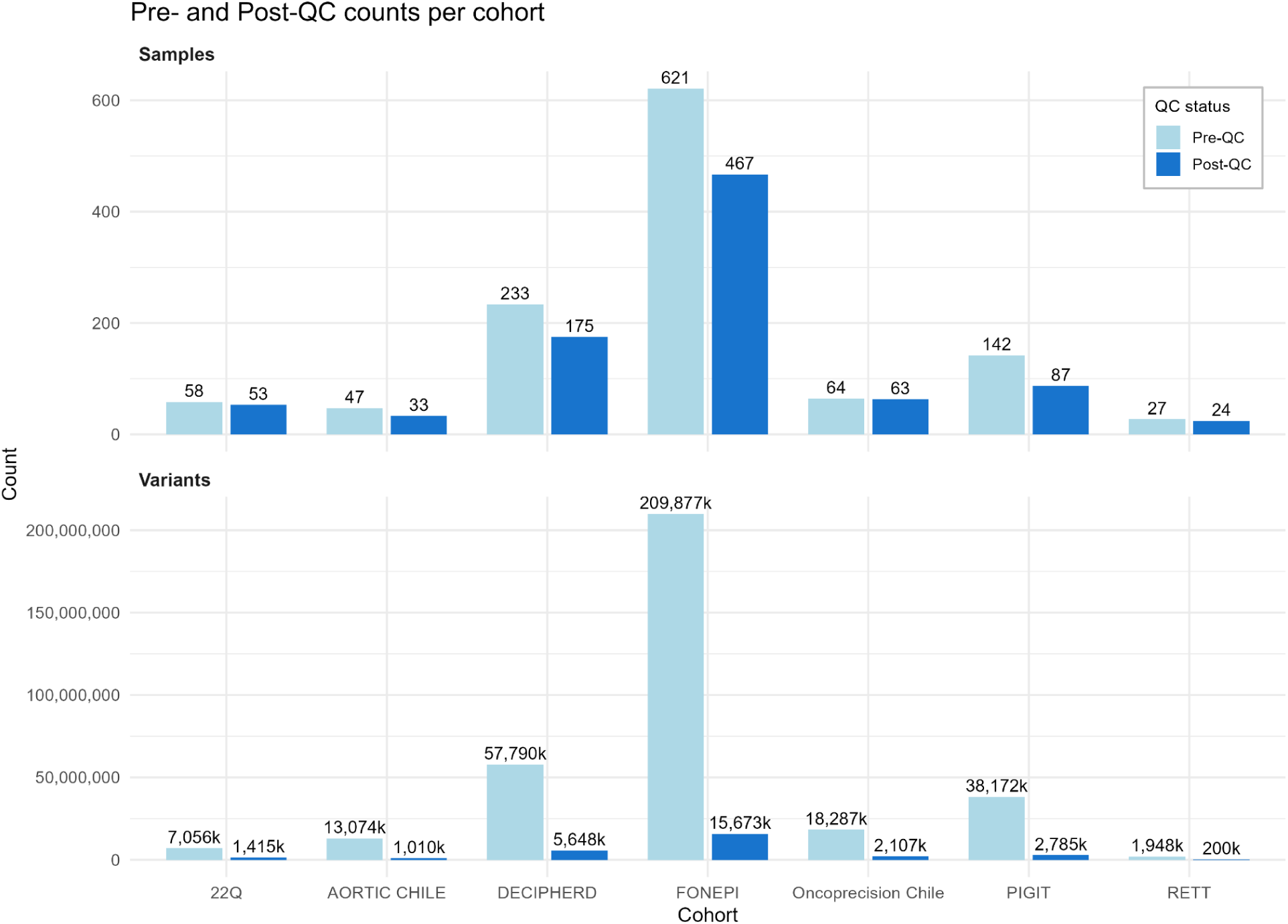
Pre- and post-quality control (QC) counts of samples and variants across the seven cohorts aggregated in CHANGER. Bars show the number of samples (top panel) and variants (bottom panel) before (light blue) and after (dark blue) QC filtering, by cohort (x-axis). Variant counts are displayed on a thousands scale.

**Supplementary Figure 2.**
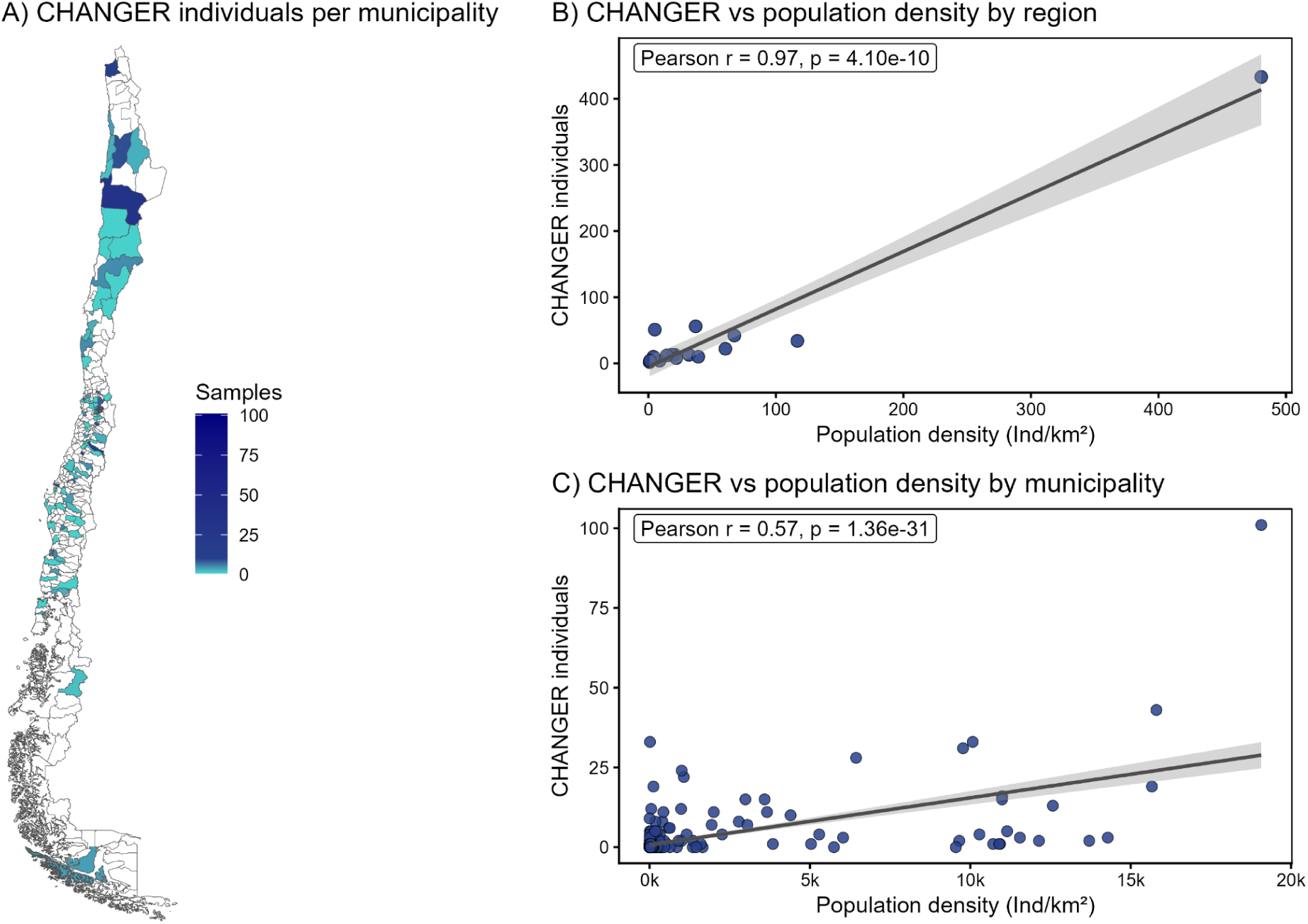
CHANGER individuals over Chile. (A) Map of Chile showing CHANGER aggregated individuals per municipality. (B) Scatterplot of CHANGER individuals versus population density by region (density in individuals per km²). (C) Scatterplot of CHANGER individuals versus population density by municipality. Data sources: polygons from the chilemapas package v3 (https://pacha.dev/chilemapas/) and population density from INE Censo 2024. (https://censo2024.ine.gob.cl/estadisticas/).

**Supplementary Figure 3.**
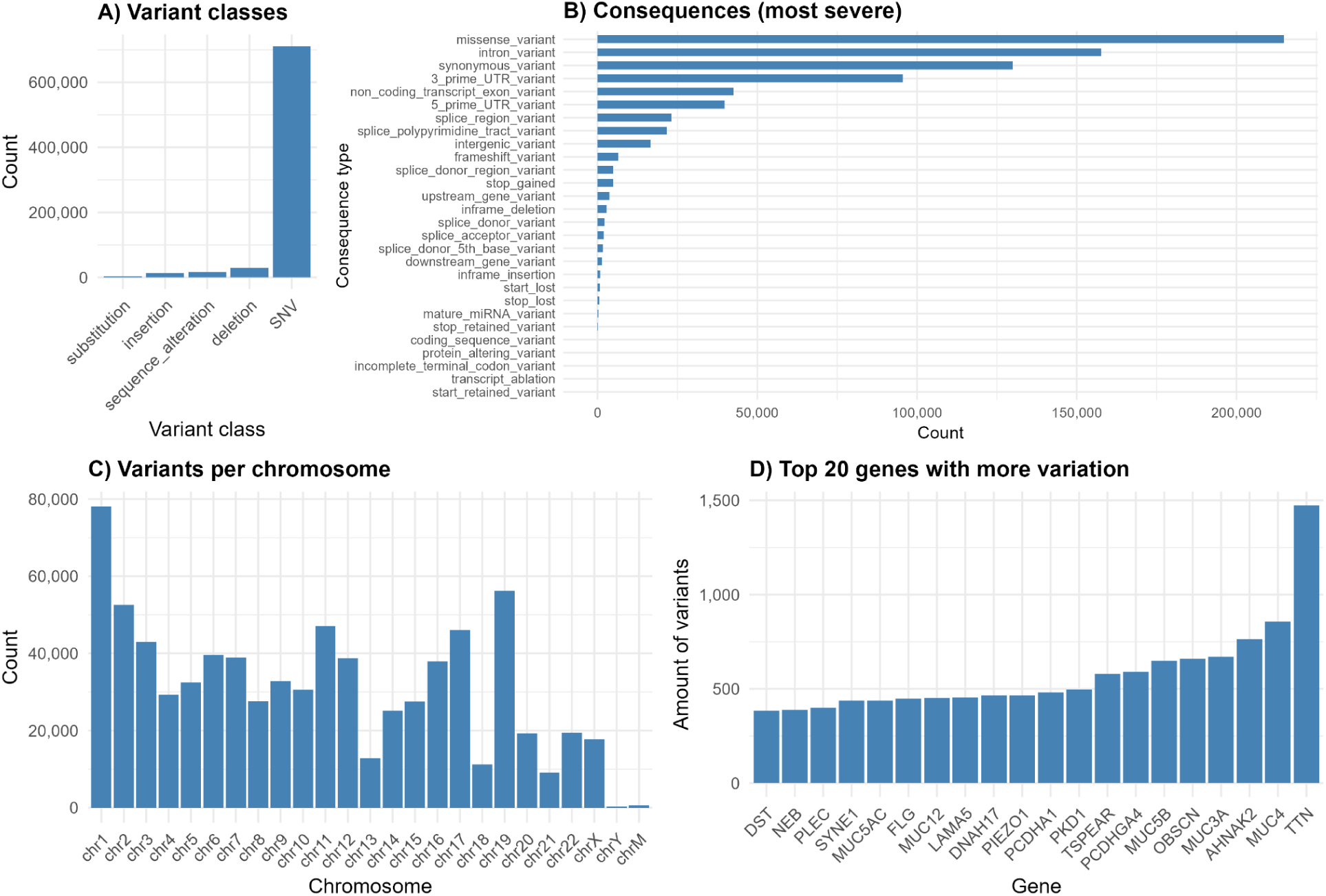
Summary of VEP (GRCh38) annotation for CHANGER variants. (A) Distribution of variant classes (SNV, deletions, insertions, substitutions, and sequence_alterations) as reported by VEP from the site-only VCF. (B) Counts of the *most severe* VEP consequence per variant, using the picked transcript (prioritized by MANE/clinical, canonical, and biotype), including coding, splice, UTR, intronic, upstream/downstream and intergenic categories. (C) Number of variants per chromosome across autosomes, sex chromosomes and mitochondrial DNA (GRCh38). (D) Top 20 genes with the highest number of distinct variants in the dataset. All annotations were generated with Ensembl VEP (offline, GRCh38 cache) using the options listed, including LOFTEE, REVEL, AlphaMissense, EVE and CADD plugins for extended functional prediction.

**Supplementary Figure 4.**
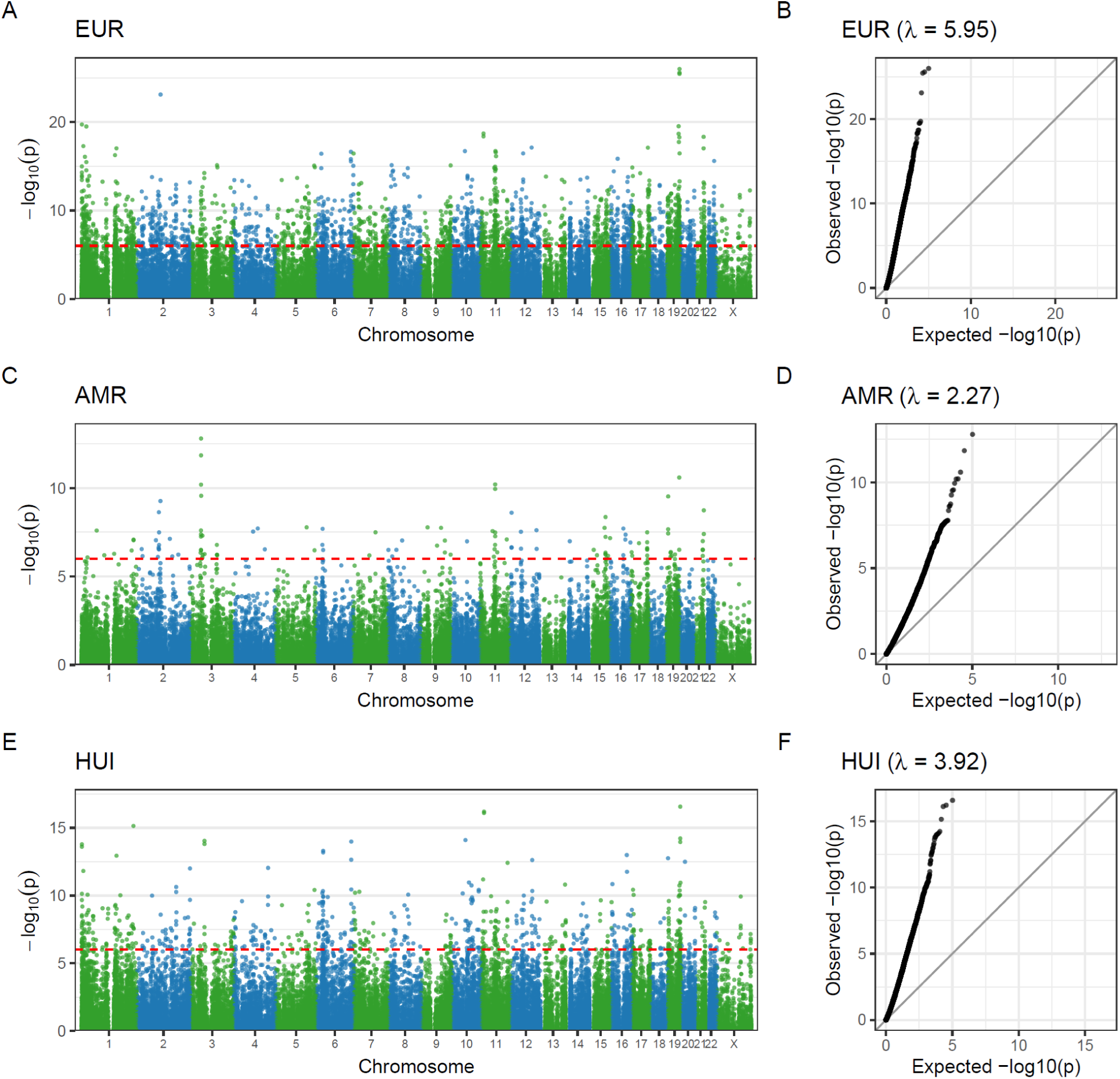
Exome-wide haplotype association with ancestry components. (A) Manhattan plot of haplotype–EUR ancestry association, showing –log10(p) for each haplotype across chromosomes; the red dashed line marks the Bonferroni-corrected significance threshold (p < 9.77×10⁻⁷). (B) QQ plot for EUR ancestry, comparing observed vs expected –log10(p) values, with genomic inflation factor λ shown in the panel title. (C–D) Equivalent Manhattan (C) and QQ (D) plots for AMR ancestry. (E–F) Equivalent Manhattan (E) and QQ (F) plots for HUI ancestry.

**Supplementary Figure 5.**
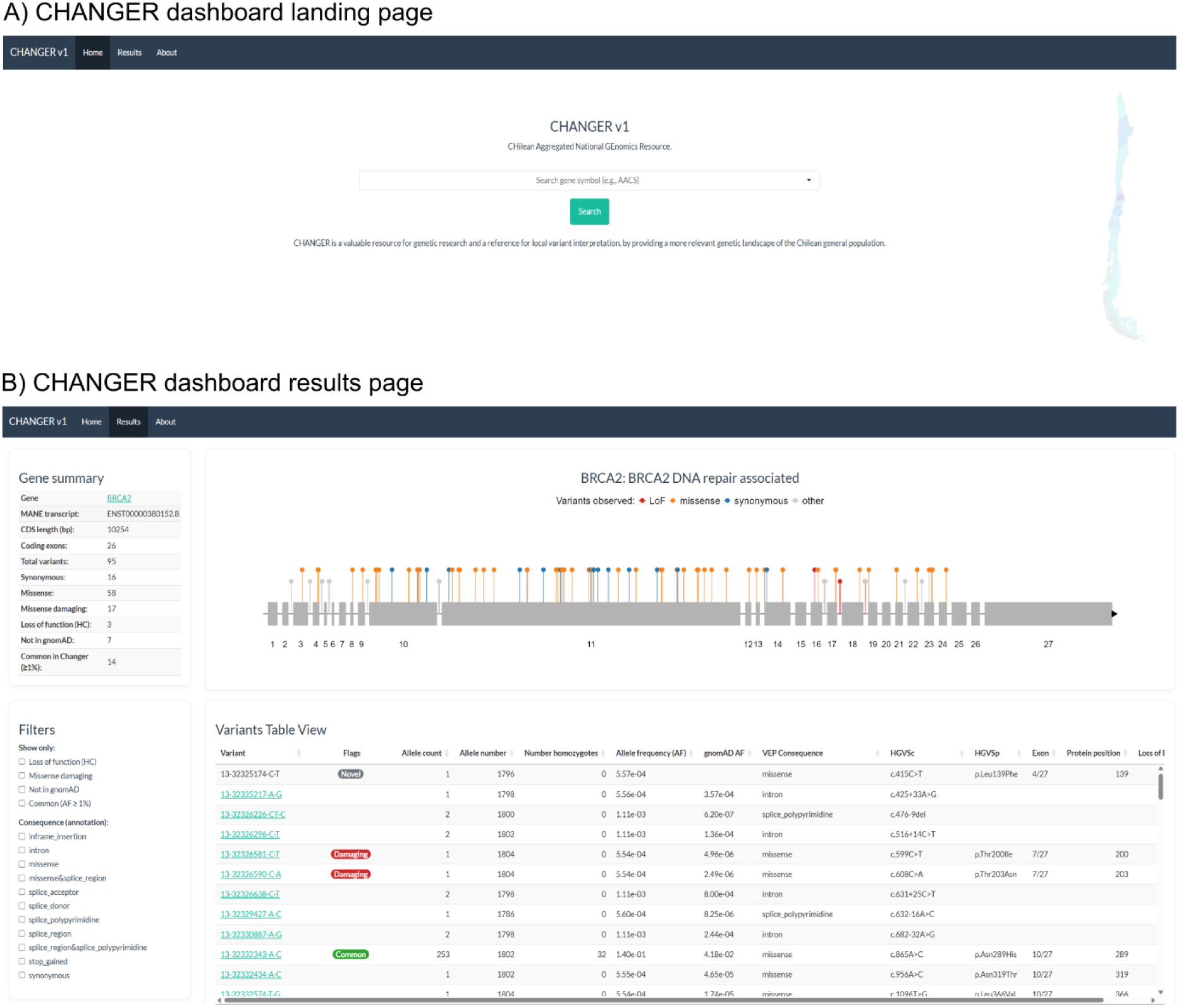
Dashboard main features. CHANGER Dashboard landing page is shown. A) A dropdown menu is available for the user to select and explore gene-wise CHANGER variants. B) CHANGER Dashboard result section consists of 4 panels: First. A gene summary panel with selected gene main features and counts. Second, A lollipop plot of observed variants mapped into the MANE transcript (introns are not proportional to length). Third, a filtering checkbox panel. Fourth, a variant table view for selected variants with featured annotations.

## References

1. 1000 Genomes Project Consortium, Auton A, Brooks LD, Durbin RM, Garrison EP, Kang HM, et al. A global reference for human genetic variation. Nature. 2015 Oct 1;526(7571):68–74.

2. Koenig Z, Yohannes MT, Nkambule LL, Zhao X, Goodrich JK, Kim HA, et al. A harmonized public resource of deeply sequenced diverse human genomes. Genome Res. 2024 Jan 5;34(5):796–809.

3. Karczewski KJ, Francioli LC, Tiao G, Cummings BB, Alföldi J, Wang Q, et al. The mutational constraint spectrum quantified from variation in 141,456 humans. Nature. 2020 May;581(7809):434–43.

4. Richards S, Aziz N, Bale S, Bick D, Das S, Gastier-Foster J, et al. Standards and guidelines for the interpretation of sequence variants: a joint consensus recommendation of the American College of Medical Genetics and Genomics and the Association for Molecular Pathology. Genet Med. 2015 May;17(5):405–23.

5. Martin AR, Kanai M, Kamatani Y, Okada Y, Neale BM, Daly MJ. Clinical use of current polygenic risk scores may exacerbate health disparities. Nat Genet. 2019 Apr;51(4):584–91.

6. Martin AR, Gignoux CR, Walters RK, Wojcik GL, Neale BM, Gravel S, et al. Human Demographic History Impacts Genetic Risk Prediction across Diverse Populations. Am J Hum Genet. 2017 Apr 6;100(4):635–49.

7. Petrovski S, Goldstein DB. Unequal representation of genetic variation across ancestry groups creates healthcare inequality in the application of precision medicine. Genome Biol. 2016 July 14;17(1):157.

8. Whiffin N, Minikel E, Walsh R, O’Donnell-Luria AH, Karczewski K, Ing AY, et al. Using high-resolution variant frequencies to empower clinical genome interpretation. Genet Med. 2017 Oct;19(10):1151–8.

9. Manrai AK, Funke BH, Rehm HL, Olesen MS, Maron BA, Szolovits P, et al. Genetic Misdiagnoses and the Potential for Health Disparities. N Engl J Med. 2016 Aug 18;375(7):655–65.

10. Wright CF, Campbell P, Eberhardt RY, Aitken S, Perrett D, Brent S, et al. Genomic Diagnosis of Rare Pediatric Disease in the United Kingdom and Ireland. N Engl J Med [Internet]. 2023 Apr 27 [cited 2025 Sept 30]; Available from: https://www.nejm.org/doi/full/10.1056/NEJMoa2209046

11. Gonzaga-Jauregui C, Gamble CN, Yuan B, Penney S, Jhangiani S, Muzny DM, et al. Mutations in COL27A1 cause Steel syndrome and suggest a founder mutation effect in the Puerto Rican population. Eur J Hum Genet. 2015 Mar;23(3):342–6.

12. Bustos BI, Pérez-Palma E, Buch S, Azócar L, Riveras E, Ugarte GD, et al. Variants in ABCG8 and TRAF3 genes confer risk for gallstone disease in admixed Latinos with Mapuche Native American ancestry. Sci Rep. 2019 Jan 28;9(1):1–12.

13. Gudmundsson S, Singer-Berk M, Stenton SL, Goodrich JK, Wilson MW, Einson J, et al. Exploring penetrance of clinically relevant variants in over 800,000 humans from the Genome Aggregation Database. Nat Commun. 2025 Oct 31;16(1):9623.

14. Lopes-Cendes I, de Oliveira TC. Inequalities and Inclusion in Genomics Applied to Healthcare: A Latin American Perspective. 2025 Jan 30 [cited 2025 June 29]; Available from: https://www.annualreviews.org/content/journals/10.1146/annurev-genom-111224-100329

15. Atkinson EG, Maihofer AX, Kanai M, Martin AR, Karczewski KJ, Santoro ML, et al. Tractor uses local ancestry to enable the inclusion of admixed individuals in GWAS and to boost power. Nat Genet. 2021 Feb;53(2):195–204.

16. Sohail M, Palma-Martínez MJ, Chong AY, Quinto-Cortés CD, Barberena-Jonas C, Medina-Muñoz SG, et al. Mexican Biobank advances population and medical genomics of diverse ancestries. Nature. 2023 Oct;622(7984):775–83.

17. Ziyatdinov A, Torres J, Alegre-Díaz J, Backman J, Mbatchou J, Turner M, et al. Genotyping, sequencing and analysis of 140,000 adults from Mexico City. Nature. 2023 Oct;622(7984):784–93.

18. Rocha CS, Secolin R, Rodrigues MR, Carvalho BS, Lopes-Cendes I. The Brazilian Initiative on Precision Medicine (BIPMed): fostering genomic data-sharing of underrepresented populations. Npj Genomic Med. 2020 Oct 2;5(1):42.

19. Naslavsky MS, Yamamoto GL, de Almeida TF, Ezquina SAM, Sunaga DY, Pho N, et al. Exomic variants of an elderly cohort of Brazilians in the ABraOM database. Hum Mutat. 2017;38(7):751–63.

20. Ministério da Saúde [Internet]. [cited 2025 Dec 5]. Genomas Brasil. Available from: https://www.gov.br/saude/pt-br/composicao/sectics/decit/genomas-brasil/genomas-brasil

21. Velasco HM, Bertoli-Avella A, Jaramillo CJ, Cardona DS, González LA, Vanegas MN, et al. Facing the challenges to shorten the diagnostic odyssey: first Whole Genome Sequencing experience of a Colombian cohort with suspected rare diseases. Eur J Hum Genet. 2024 Oct;32(10):1327–37.

22. Mariño-Ramírez L, Sharma S, Hamilton JM, Nguyen TL, Gupta S, Natarajan AV, et al. The Consortium for Genomic Diversity, Ancestry, and Health in Colombia (CÓDIGO): building local capacity in genomics and bioinformatics. Commun Biol. 2025 July 17;8(1):1062.

23. Vidal EA, Moyano TC, Bustos BI, Pérez-Palma E, Moraga C, Riveras E, et al. Whole Genome Sequence, Variant Discovery and Annotation in Mapuche-Huilliche Native South Americans. Sci Rep. 2019 14;9(1):2132.

24. COVID19 y genética del hospedero | Chilegenomico [Internet]. [cited 2025 Sept 26]. Available from: http://chilegenomico.med.uchile.cl/covid19-y-genetica-del-hospedero/

25. Eyheramendy S, Martinez FI, Manevy F, Vial C, Repetto GM. Genetic structure characterization of Chileans reflects historical immigration patterns. Nat Commun. 2015 Mar 17;6(1):6472.

26. Zhao Y, Wang Y, Shi L, McDonald-McGinn DM, Crowley TB, McGinn DE, et al. Chromatin regulators in the TBX1 network confer risk for conotruncal heart defects in 22q11.2DS. Npj Genomic Med. 2023 July 18;8(1):17.

27. Jimenez Y, Paulsen C, Turner E, Iturra S, Cuevas O, Lay-son G, et al. Exome Sequencing Identifies Genetic Variants Associated with Extreme Manifestations of the Cardiovascular Phenotype in Marfan Syndrome. Genes. 2022 June;13(6):1027.

28. Decoding complex inherited phenotypes in rare disorders: the DECIPHERD initiative for rare undiagnosed diseases in Chile | European Journal of Human Genetics [Internet]. [cited 2025 Sept 26]. Available from: https://www.nature.com/articles/s41431-023-01523-5

29. Brito F, Lagos C, Cubillos J, Orellana J, Gajardo M, Böhme D, et al. Genomic analysis in Chilean patients with suspected Rett syndrome: keep a broad differential diagnosis. Front Genet [Internet]. 2024 Mar 18 [cited 2025 Dec 5];15. Available from: https://www.frontiersin.org/journals/genetics/articles/10.3389/fgene.2024.1278198/full

30. Sealock JM, Ivankovic F, Liao C, Chen S, Churchhouse C, Karczewski KJ, et al. Tutorial: guidelines for quality filtering of whole-exome and whole-genome sequencing data for population-scale association analyses. Nat Protoc. 2025 Mar 28;1–11.

31. McKenna A, Hanna M, Banks E, Sivachenko A, Cibulskis K, Kernytsky A, et al. The Genome Analysis Toolkit: a MapReduce framework for analyzing next-generation DNA sequencing data. Genome Res. 2010 Sept;20(9):1297–303.

32. Manichaikul A, Mychaleckyj JC, Rich SS, Daly K, Sale M, Chen WM. Robust relationship inference in genome-wide association studies. Bioinforma Oxf Engl. 2010 Nov 15;26(22):2867–73.

33. Leal TP, Waldo E, Duarte-Zambrano F, Inca-Martinez M, Ramchandra J, Chaparro-Solano HM, et al. Genotype-phenotype association study conducted on LARGE-PD reveals novel loci associated with Parkinson’s Disease [Internet]. medRxiv; 2025 [cited 2025 Sept 30]. p. 2025.07.18.25331793. Available from: https://www.medrxiv.org/content/10.1101/2025.07.18.25331793v1

34. Alexander DH, Novembre J, Lange K. Fast model-based estimation of ancestry in unrelated individuals. Genome Res. 2009 Jan 9;19(9):1655–64.

35. Shriner D, Bentley AR, Gouveia MH, Heuston EF, Doumatey AP, Chen G, et al. Universal genome-wide association studies: Powerful joint ancestry and association testing. Hum Genet Genomics Adv [Internet]. 2023 Oct 12 [cited 2025 Oct 9];4(4). Available from: https://www.cell.com/hgg-advances/abstract/S2666-2477(23)00067-2

36. Frazer J, Notin P, Dias M, Gomez A, Min JK, Brock K, et al. Disease variant prediction with deep generative models of evolutionary data. Nature. 2021 Nov;599(7883):91–5.

37. Cheng J, Novati G, Pan J, Bycroft C, Žemgulytė A, Applebaum T, et al. Accurate proteome-wide missense variant effect prediction with AlphaMissense. Science. 2023 Sept 19;381(6664):eadg7492.

38. Ioannidis NM, Rothstein JH, Pejaver V, Middha S, McDonnell SK, Baheti S, et al. REVEL: An Ensemble Method for Predicting the Pathogenicity of Rare Missense Variants. Am J Hum Genet. 2016 Oct 6;99(4):877–85.

39. Schubach M, Maass T, Nazaretyan L, Röner S, Kircher M. CADD v1.7: using protein language models, regulatory CNNs and other nucleotide-level scores to improve genome-wide variant predictions. Nucleic Acids Res. 2024 Jan 5;52(D1):D1143–54.

40. Li Q, Wang K. InterVar: Clinical Interpretation of Genetic Variants by the 2015 ACMG-AMP Guidelines. Am J Hum Genet. 2017 Feb 2;100(2):267–80.

41. Fuentes Fajardo KV, Adams D, Program NCS, Mason CE, Sincan M, Tifft C, et al. Detecting false-positive signals in exome sequencing. Hum Mutat. 2012;33(4):609–13.

42. Chang CC, Chow CC, Tellier LC, Vattikuti S, Purcell SM, Lee JJ. Second-generation PLINK: rising to the challenge of larger and richer datasets. GigaScience. 2015;4:7.

43. Accurate rare variant phasing of whole-genome and whole-exome sequencing data in the UK Biobank | Nature Genetics [Internet]. [cited 2025 Dec 5]. Available from: https://www.nature.com/articles/s41588-023-01415-w

44. Mudge JM, Carbonell-Sala S, Diekhans M, Martinez JG, Hunt T, Jungreis I, et al. GENCODE 2025: reference gene annotation for human and mouse. Nucleic Acids Res. 2025 Jan 6;53(D1):D966–75.

45. Pérez-Palma E, May P, Iqbal S, Niestroj LM, Du J, Heyne HO, et al. Identification of pathogenic variant enriched regions across genes and gene families. Genome Res. 2020;30(1):62–71.

46. Pérez-Palma E, Gramm M, Nürnberg P, May P, Lal D. Simple ClinVar: an interactive web server to explore and retrieve gene and disease variants aggregated in ClinVar database. Nucleic Acids Res. 2019 July 2;47(W1):W99–105.

47. Perez G, Barber GP, Benet-Pages A, Casper J, Clawson H, Diekhans M, et al. The UCSC Genome Browser database: 2025 update. Nucleic Acids Res. 2025 Jan 6;53(D1):D1243–9.

48. INE. Instituto Nacional de Estadisticas (INE), Informe Estadísticas Vitales [Internet]. 2017 [cited 2020 Nov 20]. Available from: http://www.ine.cl/estadisticas/sociales/demografia-y-vitales/nacimientos-matrimonios-y-defunciones

49. Exome sequencing and analysis of 454,787 UK Biobank participants | Nature [Internet]. [cited 2025 Sept 30]. Available from: https://www.nature.com/articles/s41586-021-04103-z

50. Bruno LP, Doddato G, Valentino F, Baldassarri M, Tita R, Fallerini C, et al. New Candidates for Autism/Intellectual Disability Identified by Whole-Exome Sequencing. Int J Mol Sci. 2021 Jan;22(24):13439.

51. Alazami AM, Patel N, Shamseldin HE, Anazi S, Al-Dosari MS, Alzahrani F, et al. Accelerating Novel Candidate Gene Discovery in Neurogenetic Disorders via Whole-Exome Sequencing of Prescreened Multiplex Consanguineous Families. Cell Rep. 2015 Jan 13;10(2):148–61.

52. Rosamilia MB, Markunas AM, Kishnani PS, Landstrom AP. Underrepresentation of Diverse Ancestries Drives Uncertainty in Genetic Variants Found in Cardiomyopathy-Associated Genes. JACC Adv. 2023 Dec 15;3(2):100767.

53. Rodan LH, Spillmann RC, Kurata HT, Lamothe SM, Maghera J, Jamra RA, et al. Phenotypic expansion of CACNA1C-associated disorders to include isolated neurological manifestations. Genet Med Off J Am Coll Med Genet. 2021 Oct;23(10):1922–32.

54. Vesteghem C, Brøndum RF, Sønderkær M, Sommer M, Schmitz A, Bødker JS, et al. Implementing the FAIR Data Principles in precision oncology: review of supporting initiatives. Brief Bioinform. 2020 May 21;21(3):936–45.

55. Federated gnomAD | gnomAD [Internet]. [cited 2025 Dec 5]. Available from: https://gnomad.broadinstitute.org/federated

56. Best practices for data management and sharing in experimental biomedical research | Physiological Reviews | American Physiological Society [Internet]. [cited 2025 Oct 6]. Available from: https://journals.physiology.org/doi/full/10.1152/physrev.00043.2023

57. Walter K, Min JL, Huang J, Crooks L, Memari Y, McCarthy S, et al. The UK10K project identifies rare variants in health and disease. Nature. 2015 Oct;526(7571):82–90.

58. McCarthy S, Das S, Kretzschmar W, Delaneau O, Wood AR, Teumer A, et al. A reference panel of 64,976 haplotypes for genotype imputation. Nat Genet. 2016;48(10):1279–83.

59. Taliun D, Harris DN, Kessler MD, Carlson J, Szpiech ZA, Torres R, et al. Sequencing of 53,831 diverse genomes from the NHLBI TOPMed Program. Nature. 2021 Feb;590(7845):290–9.

60. Nacional B del C. 2015 [cited 2021 Aug 22]. Biblioteca del Congreso Nacional | Ley Chile. Available from: https://www.bcn.cl/leychile

61. Valdés E, Lecaros JA. Biobanks and data interoperability in Latin America: engendering high-quality evidence for the global research ecosystem. Front Med [Internet]. 2024 Dec 16 [cited 2025 Dec 5];11. Available from: https://www.frontiersin.org/journals/medicine/articles/10.3389/fmed.2024.1481891/full

